# Age related clustering in global COVID-19 infection fatality ratios and death trajectories

**DOI:** 10.1101/2020.08.11.20172478

**Authors:** Thu-Lan Kelly, Caroline Miller, Jacqueline A Bowden, Joanne Dono, Paddy A Phillips

## Abstract

**Background:** An accurate measure of the impact of COVID-19 is the infection fatality ratio, or the proportion of deaths among those infected, which is independent of variable testing rates between nations. The risk of mortality from COVID-19 depends strongly on age and current single estimates of the infection fatality ratio do not account for differences in national age profiles. In addition, it is unclear whether age influences cumulative death trajectories, or if differences between regions are because of the effect and timing of public health interventions.

Our objective is to determine whether (1) infection fatality ratios and (2) death trajectories are clustered into more than one group due to differences in national age profiles.

**Methods:** National age standardised infection fatality ratios were derived from age stratified estimates from China and population estimates from the World Health Organisation. The infection fatality ratios were clustered into groups using Gaussian mixture models. Trajectory analysis clustered cumulative death rates at two time points, 50 and 150 days after the first reported death.

**Findings:** Infection fatality ratios from 201 nations were clustered into three groups: young, middle and older, with corresponding means (SD) of 0.20% (0.03%), 0.38% (0.11%) and 0.93% (0.21%). At 50 and 150 days, there were two and three clusters, respectively, of cumulative death trajectories from 122 nations with at least 25 deaths reported at 100 days. The first cluster had steadily increasing or stable cumulative death rates, while the second and third clusters had moderate and fast increases in rates, respectively. Fifty-eight nations changed cluster group membership between time points. There was an association between the infection fatality ratio clusters and the change in trajectory clusters between 50 and 150 days (p=0.014).

**Conclusion:** Differences in national age profiles created three clusters in the COVID-19 infection fatality ratio, with the impact on younger nations less than the current estimate 0.5-1.0%. National cumulative death rates were clustered into steady, moderate or fast trajectories. Changes in death rate trajectories between 50 and 150 days were associated with the infection fatality ratio clusters, however evidence for the influence of age on death trajectories is mixed.

## Introduction

The disease COVID-19 caused by the coronavirus SARS-CoV-2 was first described in Wuhan, China in December 2019 [1-2]. The impact the disease will have on the global population has been estimated through the case fatality rate (CFR), or the proportion of deaths among confirmed cases. Since the number of cases depends on testing rates which may include only symptomatic or severe cases, the CFR may be an over-estimate of the impact of the disease.

In contrast, the infection fatality ratio (IFR) is the proportion of deaths among infected individuals and is a more accurate estimate of disease mortality. However, it is difficult to determine the true number of infections in a population. Recent antibody prevalence studies have attempted to establish infection rates in the USA, Spain and elsewhere [3-5].

Studies have shown that the risk of death from COVID-19 depends strongly on age [6-10]. A recent meta-analysis calculated an IFR of 0.68 % (95% CI 0.53-0.82%), but with significant heterogeneity between regions [6]. The authors concluded that different regions may experience different IFRs due to age structure and underlying co-morbidities and called for more research on age stratified IFRs. A further meta-analysis on the age specificity of COVID-19 IFRs [7] confirmed the exponential dependence of mortality on age also found in studies from China [8] and Italy [9]. Overall IFRs varied from 0.66% (95% credible interval 0.39-1.33%) in China to 1.29 % (95% crI 0.89-2.01%) in Italy, because Italy has an older population than China. Differences in age structure and age specific prevalence were found to account for up to 90% of the geographic variation in population IFR [7] and an IFR of below 0.5% was ruled out in populations with more than 30% over 60 years old [9]. Due to this strong age dependence, the US Centre for Disease Control and Prevention now publishes age-specific estimates of IFR [10].

The IFR summarises the expected mortality risk in a population at a single point in time. In addition to the IFR, the rate at which the disease spreads throughout a population and the death rate also has a significant impact on health resources. While age is a significant risk factor for mortality, the spread of infection also depends on public health interventions such as physical distancing measures, mask wearing, testing, contact tracing, quarantine and border controls [11]. It is unclear what effect age has on increases in COVID-19 death rates over time. Clustering groups of countries with similar cumulative death rate trajectories at different time points enables comparisons of the timing and effectiveness of mitigation strategies. Comparisons of death trajectory clusters with and without adjusting for differences in national age profiles may inform whether age influences the spread of the disease throughout a population.

## Methods

Age standardisation of IFRs estimated from China by Verity et al [8] was performed using a weighting method (see Supporting Material). The overall estimate of the IFR from China (0.66% [8]) was multiplied by each nation’s weight to obtain a point estimate of the age adjusted IFR.

Model based clustering with Gaussian mixture models was used to cluster groups of nations with IFRs arising from the same normal distribution, using the ‘mclust’ R package version 5.4.6 [12]. Estimates of the mean IFR, SD and bootstrapped 95% confidence intervals of the mean were determined for each distribution.

To investigate factors related to death rates that are independent of national age profiles, such as public health interventions, the next stage was to analyse death rate trajectories. If infection rates are assumed to be equal across groups and age stratified IFRs relative to those in the 80+ age group are assumed to be the same for every nation, then the IFR weights can be used to age standardise death rates per population. The age stratified IFRs in China [8] are broadly in agreement with those estimated by a meta-analysis [7] and in Italy [9] (S1 Table in the Supporting Material). To remove any potential effect of age on the death trajectories, cumulative death rates were weighted. Trajectory analysis of cumulative death rates per population was used at two time points: 50 and 150 days after the first reported death from COVID-19. Since trajectory analysis is sensitive to outliers in the data [13], to ensure stable trajectories, only countries with at least 25 deaths reported by 100 days after the first death were included in the analysis. Rolling 14-day averages of weighted cumulative deaths rates were smoothed using splines to reduce the effect of outlying data points. The R package ‘traj’ version 1.2, which combines principal components of statistical measures of growth and cluster analysis, was used to cluster cumulative death rate trajectories into groups, without requiring the number of clusters to be determined a priori [13]. A sensitivity analysis was also conducted using unweighted trajectories.

To test whether age may be associated with death rate trajectories, Fisher’s exact test was used to test for differences in the IFR cluster group membership and any change in trajectory group membership between the two time points.

The R package ‘rworldmap’ version 1.3-6 [14] was used to visualise the IFR and trajectory clusters on a global scale. R software version 4.0.2 (R foundation for Statistical Computing, Vienna, Austria) was used for all analyses.

## Data

Estimates of national populations in 2020 by five-year age groups from the World Health Organisation were available from [15]. Daily cumulative death rates compiled by the European Centre for Disease Control were obtained from the Our World in Data website [16]. All datasets and R code used to produce the results are available from https://github.com/lan-k/COVID19.

## Results

Age adjusted IFRs were calculated for 201 countries. If the national IFRs were assumed to be from one normal distribution, the mean IFR would be 0.54% (SD 0.34%); however, a histogram of the estimated IFRs showed that these may not be represented by a single normal distribution (Fig 1).

**Figure 1:**
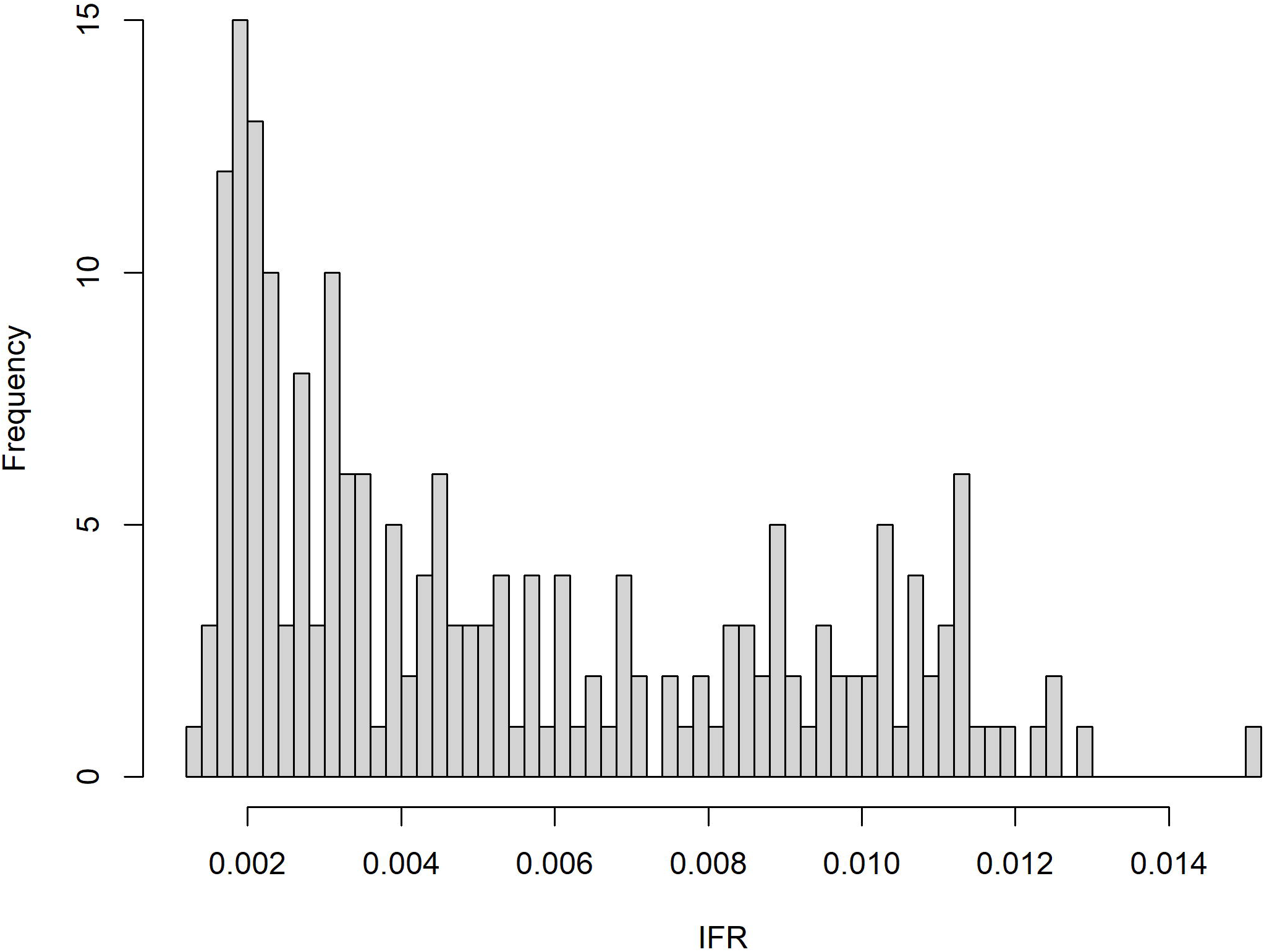
Histogram of global IFR estimates.

Clustering of the IFRs produced three groups of nations with young, middle and older age profiles. Model fit diagnostics can be found in the Supporting Material (S1 and S2 Figs). Mean IFRs (SD) from the three distributions are 0.20% (0.03%), 0.38% (0.11%) and 0.93% (0.21%) (Table 1). Bootstrapped 95% CIs for the mean and SD of the three normal distributions and the minimum and maximum IFR in each cluster are also presented in Table 1. The countries included in each cluster are displayed in Fig 2. After excluding countries in the ‘Young’ cluster, the mean IFR (95% CI) from the remaining countries, assuming the data were from a single normal distribution, was 0.67% (0.62-0.72%), which is very close to the meta-analysis estimate in middle aged and older nations of 0.68% (95% CI 0.53-0.82%) [6].

**Table 1:**
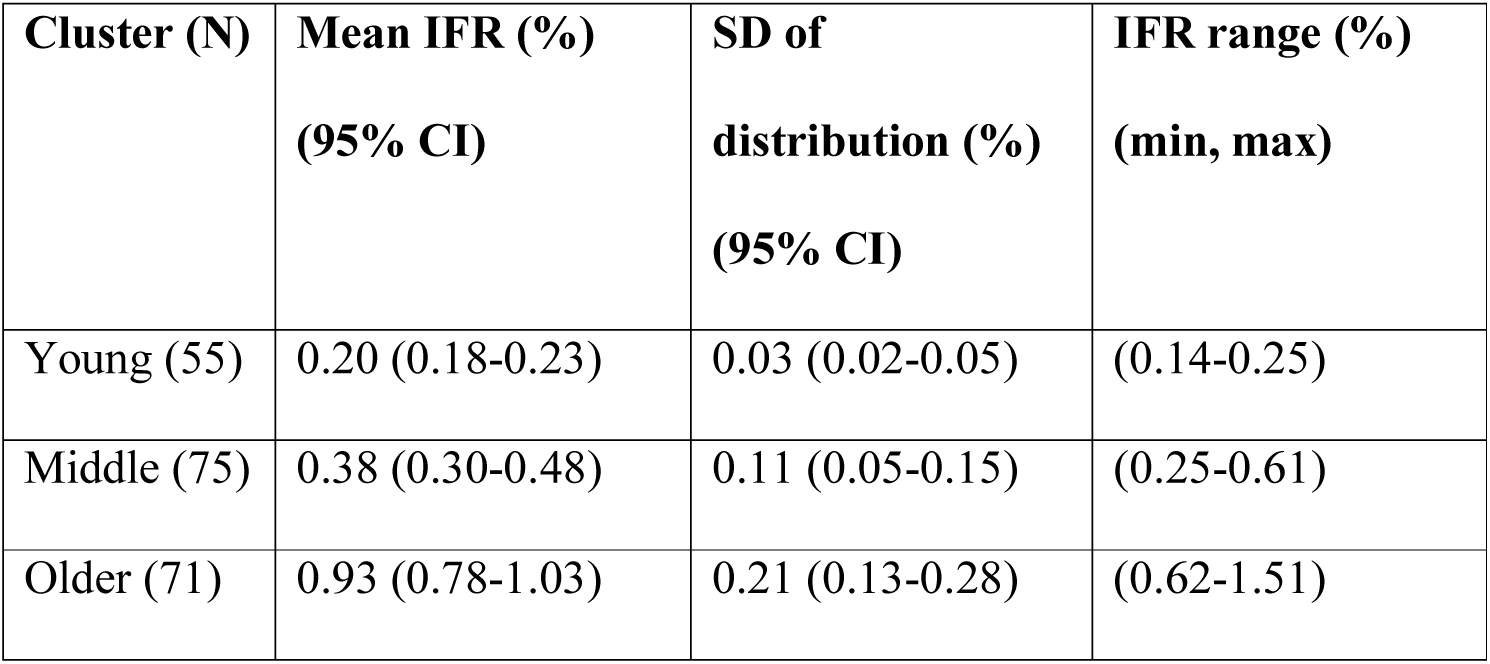
Characteristics of the three IFR clusters.

**Figure 2:**
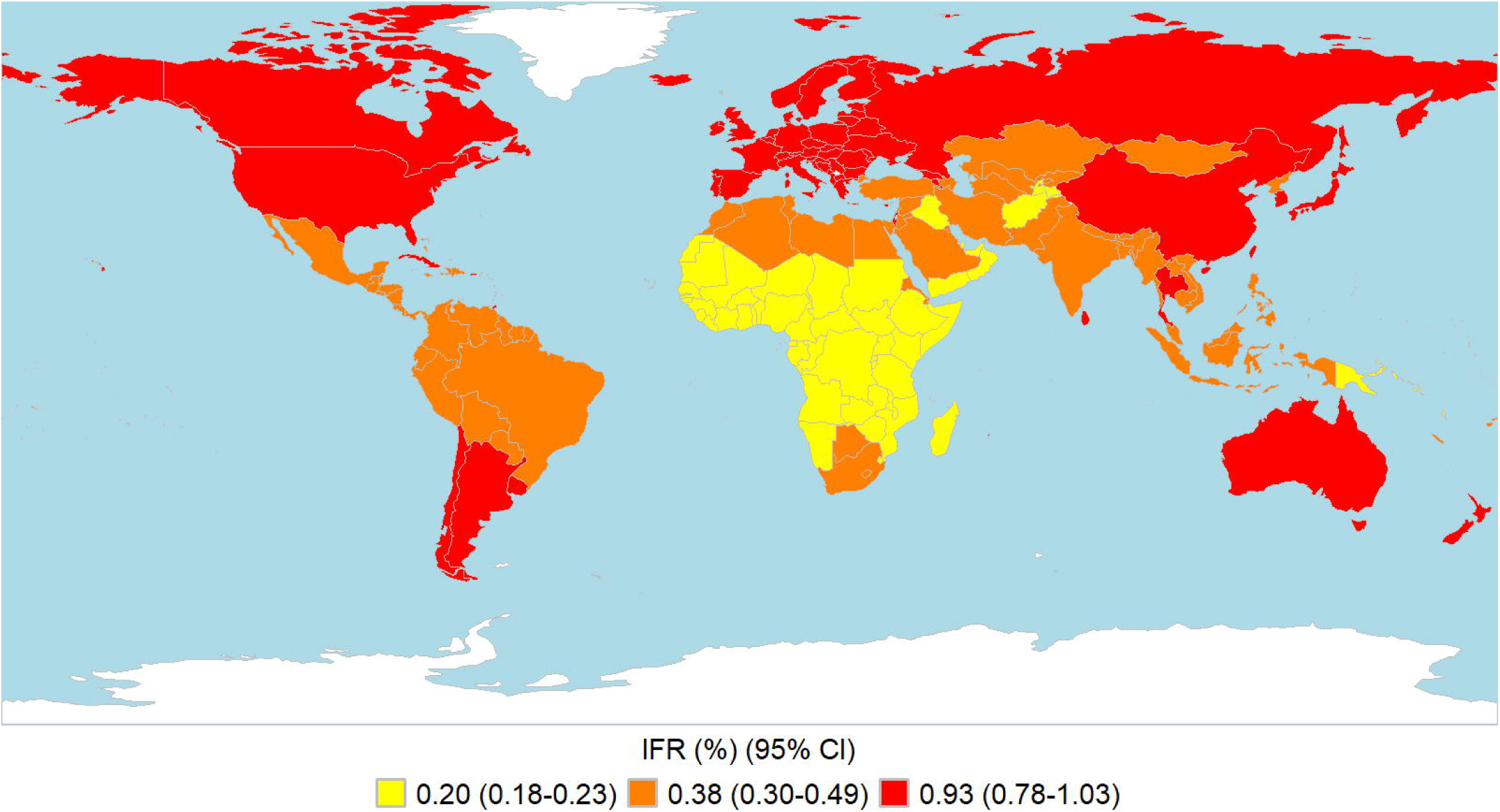
National membership of three IFR clusters. Countries with missing data are shown in white.

For the trajectory analysis, cumulative death rates were available for 122 countries with at least 25 deaths 100 days after the first reported death, as of October 13, 2020. Trajectory analysis clustered countries based on the growth of weighted cumulative death rates over time and the groups are independent of the IFR clusters in Fig 2. Two and three clusters were found at 50 and 150 days, respectively; ‘Steady’, ‘Moderate’ and a third cluster ‘Fast’ at 150 days (Table 2). Cluster group membership was based on the shape of the trajectory rather than the value of cumulative death rates at the end of the period. The first cluster, ‘Steady’, had cumulative death rates which had plateaued or slowly increased towards the end of the time window, while the second and third clusters, ‘Moderate’ and ‘Fast’, showed death rates which were moderately or rapidly increasing, respectively. Details of the statistical measures which defined the clusters at each time window and diagnostics can be found in S2 Table and S3 and S4 Figs in the Supporting Material. The median trajectories and interquartile ranges for each group at 50 and 150 days after the first reported death are shown in Fig 3. Since clusters were defined by the shape of the trajectories, cluster group membership did not change in a sensitivity analysis using unweighted trajectories. Fig 4 shows national group membership of the trajectory clusters at 50 and 150 days after the first reported death.

**Table 2:** Median cumulative death rate and interquartile range for clusters at 50 and 150 days after the first reported death.

| Cluster | 50 days<br>(N) | 150 days<br>(N) | Median cumulative<br>death rate at 50 days<br>(IQR) (per 100k) | Median cumulative<br>death rate at 150 days<br>(IQR) (per 100k) |
| --- | --- | --- | --- | --- |
| Steady | 75 | 61 | 8.63 (4.10-18.0) | 30.3 (12.3-55.0) |
| Moderate | 47 | 38 | 18.2 (3.9-48.0) | 70.6 (37.3-189) |
| Fast | - | 23 | - | 372 (295-492) |

**Figure 3:**
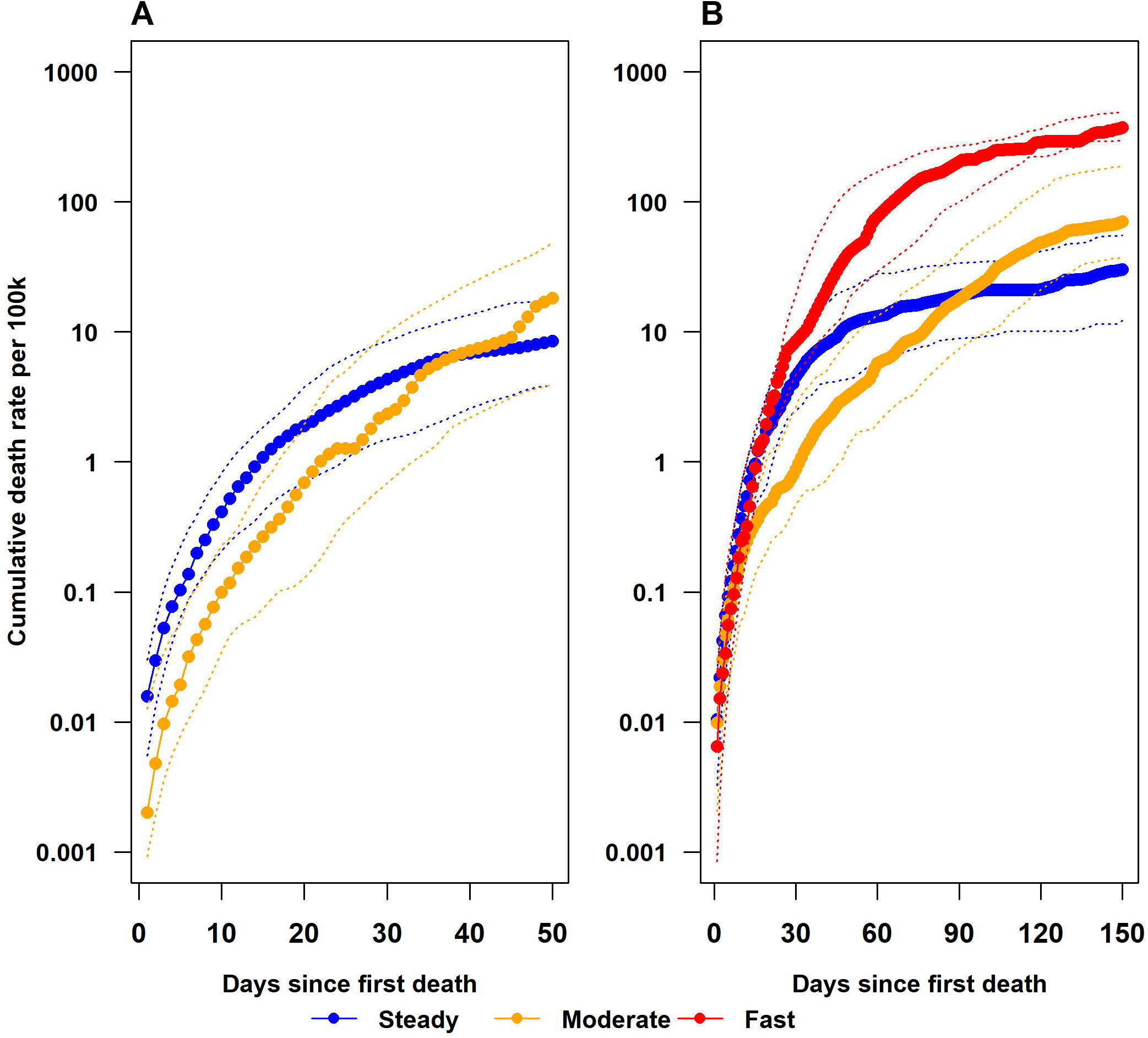
Median cluster trajectories. Median trajectories (solid line) and interquartile range (dashed line) for the ‘Steady’ (blue), ‘Moderate’ (orange) and ‘Fast’ (red) clusters at (A) 50 days and (B) 150 days after the first reported death.

**Figure 4:**
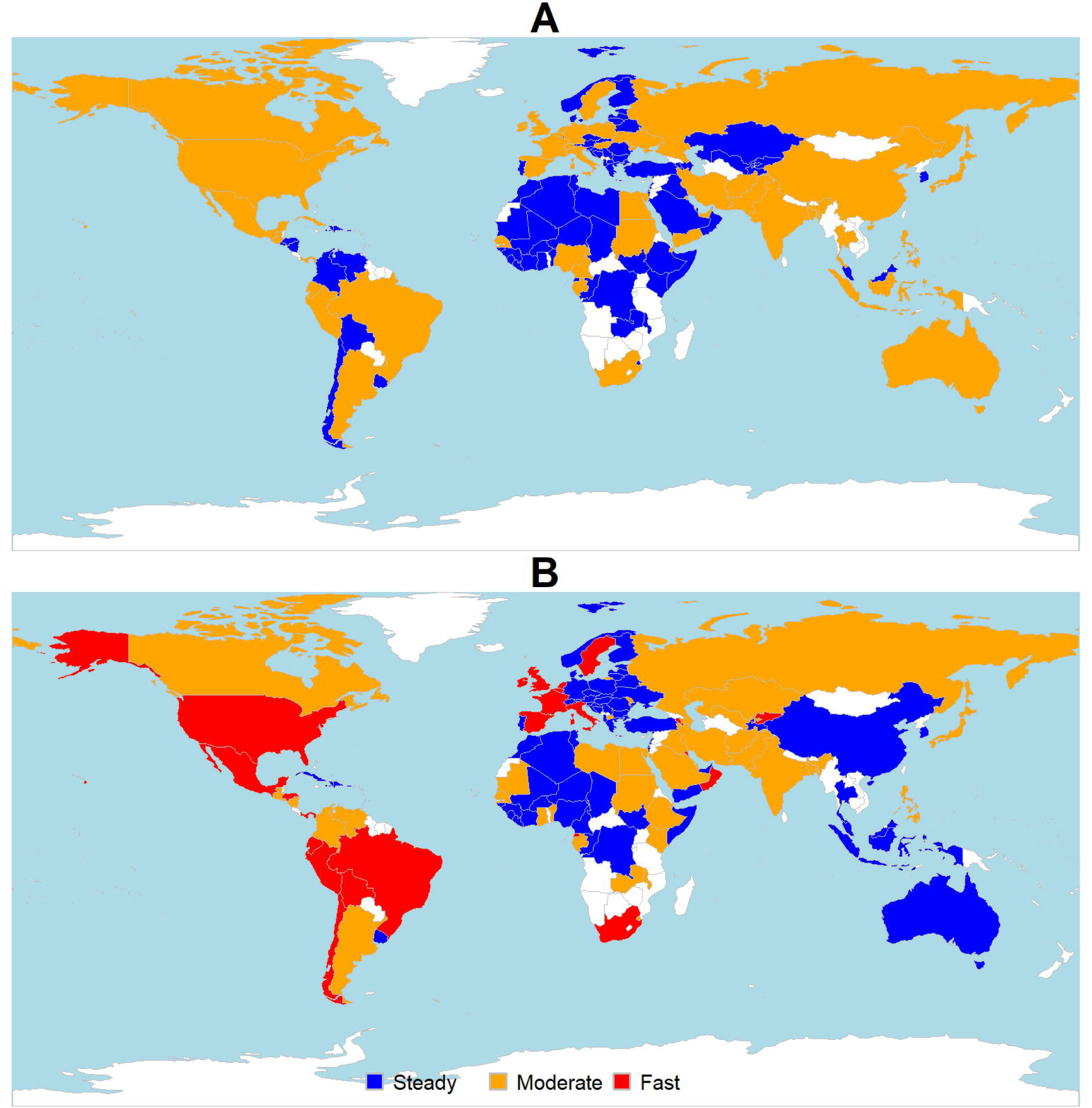
National trajectory cluster group membership. Cluster group membership of the ‘Steady’ (blue), ‘Moderate’ (orange) and ‘Fast’ (red) clusters at (A) 50 days and (B) 150 days after the first reported death. Countries excluded from the analysis are shown in white.

Between the two time points, 58 out of 122 nations (48%) changed cluster group membership (Fig 5). Forty-four countries in the ‘Worse’ group moved to faster death trajectories, either from ‘Steady’ at 50 days to ‘Moderate’ or ‘Fast’ at 150 days; or from ‘Moderate’ at 50 days to ‘Fast’ at 150 days. There were 14 countries in the ‘Improved’ group which changed from ‘Moderate’ at 50 days to ‘Steady’ at 150 days.

**Figure 5:**
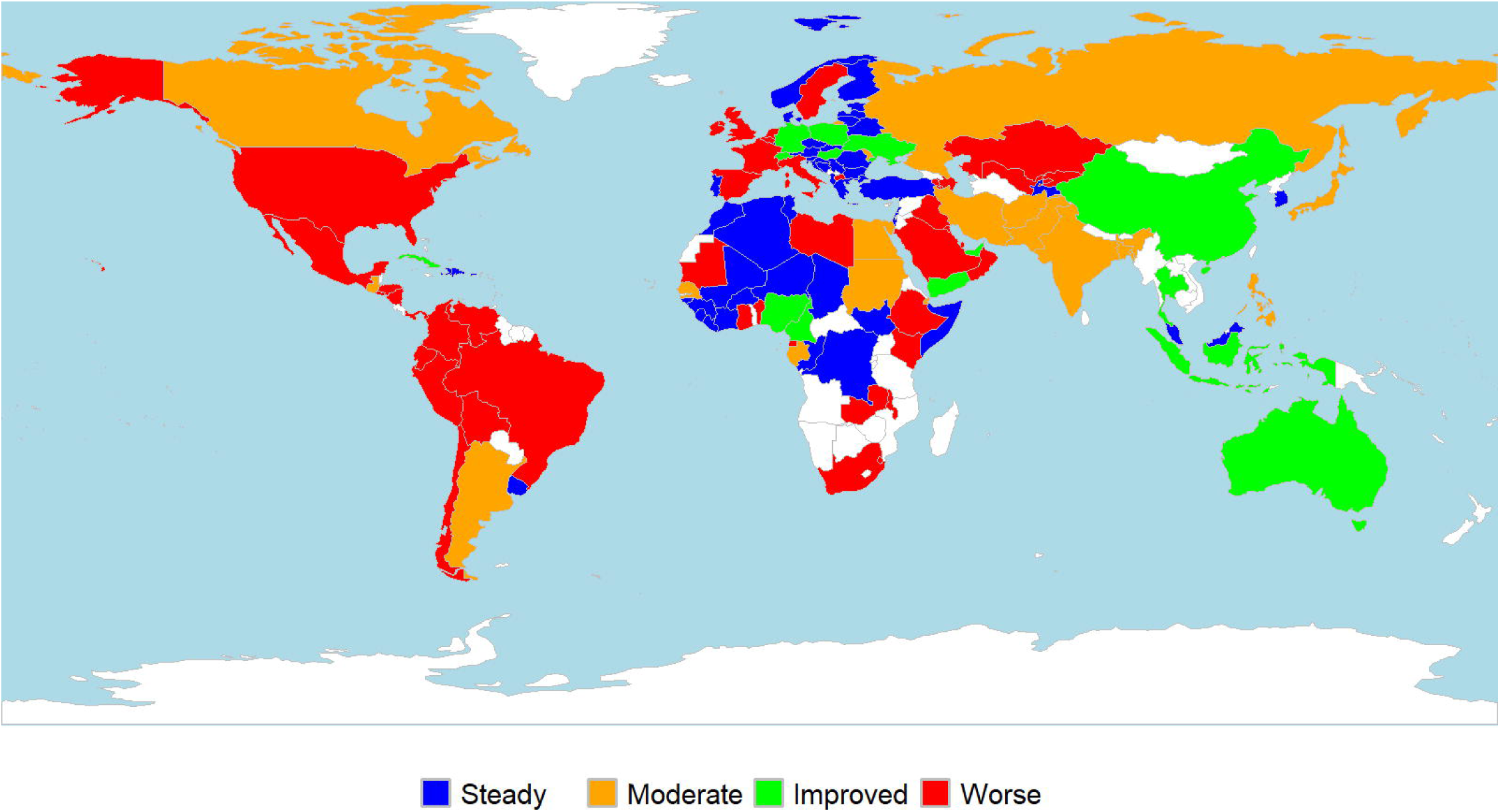
Change in trajectory cluster group membership over time. Change in group membership of trajectory clusters between 50 and 150 days after the first reported death. The ‘Steady’ (blue) and ‘Moderate’ (orange) groups remained unchanged between time points. Between 50 and 150 days, ‘Improved’ (green) changed from ‘Moderate to ‘Steady’, while ‘Worse’ (red) increased death rates to ‘Moderate’ or ‘Fast’.

Despite the trajectory clusters remaining unchanged between the weighted and unweighted analyses, there was evidence of an association between the IFR clusters and the change in cluster group membership of the death rate trajectories (Table 5, p=0.014). Middle aged countries were more likely to be ‘Worse’ between 50 and 150 days, while older nations were more likely to be ‘Steady’. Young nations showed mixed patterns of change; they were just as likely to be ‘Moderate’ as ‘Improved’, or ‘Steady’ as ‘Worse’. Compared with Table 2, there were 20/55 (36%) young, 39/75 (52%) middle and 4/71 (6%) older nations which were excluded due to low numbers of reported deaths at 100 days.

**Table 5:**
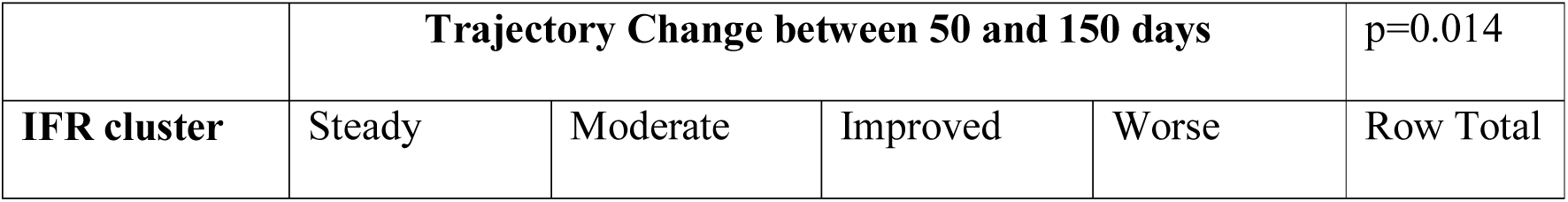
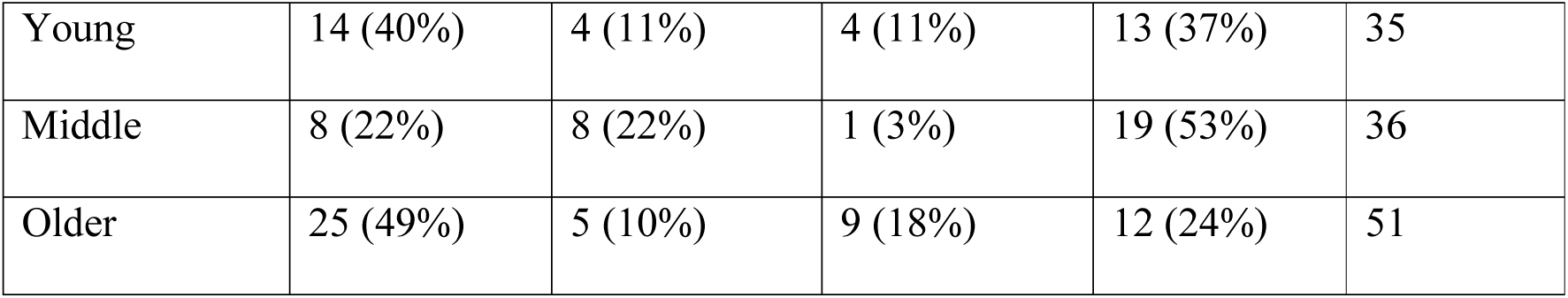
Association between the IFR clusters and the change in trajectory membership between 50 and 150 days. Cells show N and row % for 122 nations. Row percentages may not add to 100% due to rounding. P-value from Fisher’s exact test.

## Discussion

The risk of death from COVID-19 is highly dependent on age. Current estimates of the IFR are between 0.5-1.3%, are difficult to calculate and assume a single value will describe the global impact of the disease. There have been few studies reporting IFRs for younger nations such as in Africa, possibly due to difficulties in testing, measuring the number of asymptomatic infections and reporting accurate death rates. It has been suggested the different regions will experience different IFRs due to age structure and co-morbidities [6, 17-18]. A single estimate of the IFR for all nations may not capture the true global distribution. Other studies have predicted COVID-19 IFRs are reduced in low to middle income countries, even after adjusting for limited health system capacity [18-19]. Conversely, older nations experience higher IFRs, even those with more advanced health systems.

Our study has shown that national IFRs estimated using direct age standardisation of Chinese data are not drawn from a single normal distribution, but from a mixture of three distributions with different means and standard deviations. This would explain some of the heterogeneity in the IFRs reported [6]. When countries from the ‘Young’ cluster were excluded, the mean of the remaining national IFRs, assuming a single normal distribution, is very close to the meta-analysis estimate in middle aged and older nations. If data from younger countries become available, they may confirm our findings. While younger nations may have lower age standardised IFRs, these countries may have less developed health systems and poorer health status, so the actual infection fatality ratio in these nations may be higher than that estimated due to age alone [18].

While mortality risk increases exponentially with age, other factors may influence the spread of the disease and death rates over time. Trajectory analysis clustered nations depending on the growth in death rates at 50 and 150 days after the first reported death. At 50 days, North America, most of Europe and Asia, parts of Central and South America and Australia were experiencing moderately increasing death rates. By 150 days, parts of South-East Asia, Eastern Europe and Australia (the ‘Improved’ group in Fig 5) had stabilised their death trajectories through public health interventions such as lockdowns, increasing testing rates, mask wearing, contact tracing and border controls. In contrast, there were accelerating death rates in the USA, parts of Central and South America, the Middle East and Africa at 150 days (the ‘Worse’ group), while the ‘Moderate’ group included regions in South and East Asia, North Africa, Canada, Argentina and Russia which had not slowed their trajectories. The time window at 150 days may indicate that some nations were experiencing subsequent waves of infections due to the easing of restrictions or other factors, including those who had successfully suppressed the disease previously.

We found mixed evidence for the influence of age on death rate trajectories. The trajectory clusters were the same in the weighted and unweighted analyses at both time points and each trajectory cluster included a mixture of young, middle and older nations from the IFR clusters. However, we found an association between the IFR clusters and a change in the trajectory cluster group membership between 50 and 150 days. The young and middle aged IFR clusters are very similar to the low- and middle-income nations discussed in Walker et al [19], so this apparent association may be due to other factors. For example, low income (younger) nations acted earlier with suppression strategies due to limited health system capacity [19] and nations which successfully suppressed the disease had too few deaths to appear in the trajectory analysis. Apart from relatively immutable risk factors in a population, such as gender and co-morbidities [17-19], implementation of public health measures can make an important difference to the increase in deaths. Further investigation is required to determine the effect of age on death rate trajectories.

In addition to age, mortality also depends on gender, co-morbidities, ethnicity, obesity and other risk factors such as smoking [20-21], as well as access to health services. Increased risk of severe COVID-19 requiring hospitalisation due to underlying health conditions has been calculated at the national level [17]. As an extension to our analysis, the risk models developed for infection hospitalisation ratios by Clark et al [17] could be adapted to the age standardised national IFR point estimates before clustering. However, Levin et al [7] found that age profiles and age-related prevalence accounted for up to 90% of geographic variation in national IFRs.

Our study has some limitations. Age stratified IFRs relative to the 80+ age group may differ from those in China or Italy, particularly in countries where health system support is limited, overwhelmed or inequitable. The assumption that infection rates are equal across age groups may be met only in nations with large outbreaks and high death rates or with high inter-generational mixing [19, 22-23]. However, cluster group membership in the trajectory analysis did not depend on weighting for age adjustment. COVID-19 mortality data may be under-reported and the calculated IFRs may be under-estimates or lower bounds [18]. Conversely, mortality from COVID-19 may reduce throughout the pandemic as more effective treatments for the disease are discovered [24] and the calculated IFRs may become upper bounds. The IFR estimates were produced from data that was available in February 2020, before large scale seroprevalence studies had been conducted [3-6]. If more up to date age stratified IFR estimates become available, the analysis can be updated. The spread of disease through a population may also depend on international mobility, climate or regional susceptibility [25]. Finally, the association between the IFR clusters and the change in death rate trajectories between time points may be biased by socioeconomic factors or missing death rate trajectories in countries excluded due to low numbers of reported deaths.

## Conclusion

Age standardised COVID-19 IFRs were clustered into three groups depending on national age profiles. A cluster of younger nations, predominantly in Africa, had a lower mean IFR than older nations. However, these countries may have less developed health systems and poorer overall health status, so the actual IFR in these nations may be higher than that estimated from age alone. The change in death rate trajectories over time was associated with age but may also be due in part to other factors such as public health interventions.

It is important to consider the national age structure in planning for the impact of COVID-19 on overall mortality, however public health interventions are important in reducing the spread of the disease and hence death rates in a population over time.

## Data Availability

All data are fully available without restriction.

https://github.com/lan-k/COVID19

## Supporting information captions

**S1 Table: Comparison of global age specific and relative IFRs**. Confidence limits for IFR/IFR 80+ were estimated using the upper and lower limits for IFR and dividing by the point estimate for IFR 80+ (IFR 81+ for Italy).

**S1 Figure**: **Probability density function and histogram**. Probability density function of fitted distributions from three clusters (solid line) and histogram of observed IFRs.

**S2 Figure: Model diagnostics**. Quantile-Quantile plots to test normality (left) and estimated and empirical cumulative density functions (CDF) (right).

**S2 Table: Statistical measures of trajectory clusters**. Statistical measures selected by factor analysis to describe the 50 and 150 day trajectories by cluster.

**S3 Figure: Criteria used to determine optimal number of clusters for 50 day trajectories**. Cubic clustering criterion (ccc) criteria for 2-15 clusters (left) and within groups sum of squares for 1-15 clusters (right).

**S4 Figure: Criteria used to determine optimal number of clusters for 150 day trajectories**. Cubic clustering criterion (ccc) criteria for 2-15 clusters (left) and within groups sum of squares for 1-15 clusters (right).

## References

1. Zhu N, Zhang D, Wang W, et al; China Novel Coronavirus Investigating and Research Team. A novel coronavirus from patients with pneumonia in China, 2019. N Engl J Med. 2020;382(8):727–733. doi:10.1056/NEJMoa2001017

2. Novel Coronavirus Pneumonia Emergency Response Epidemiology Team. Vital surveillances: the epidemiological characteristics of an outbreak of 2019 novel coronavirus diseases (COVID-19)—China, 2020. China CDC Weekly. 2020;2(8):113-Accessed April 16, 2020. http://weekly.chinacdc.cn/en/article/id/e53946e2-c6c4-41e9-9a9b-fea8db1a8f51

3. Havers FP, Reed C, Lim T, et al. Seroprevalence of Antibodies to SARS-CoV-2 in 10 Sites in the United States, March 23-May 12, 2020. JAMA Intern Med. Published online July 21, 2020. doi:10.1001/jamainternmed.2020.4130

4. Pollán M, Pérez-Gómez B, Pastor-Barriuso R, Oteo J, Hernán MA, Pérez-Olmeda M, et al. Prevalence of SARS-CoV-2 in Spain (ENE-COVID): a nationwide, population-based seroepidemiological study. Lancet 2020; published online July 6. http://dx.doi.org/10.1016/S0140-6736(20)31483-5.

5. Stringhini S, Wisniak A, Piumatti G, Azman AS, Lauer SA, Baysson H, et al. Seroprevalence of anti-SARS-CoV-2 IgG antibodies in Geneva, Switzerland (SEROCoV-POP): a population-based study. Lancet 2020; published online June 11. http://dx.doi.org/10.1016/S0140-6736(20)31304-0

6. Meyerowitz-Katz G, Merone L, A systematic review and meta-analysis of published research data on COVID-19 infection-fatality rates. International Journal of Infectious Diseases 2020; published online, September 29. https://doi.org/10.1016/j.ijid.2020.09.1464

7. Levin AT, Hanage WP, Owusu_Boaitey N, et al. Assessing the Age Specificity of Infection Fatality Rates for COVID-19: Systematic Review, Meta-Analysis, and Public Policy Implications. Preprint published online October 8, 2020. Available at https://doi.org/10.1101/2020.07.23.20160895

8. Verity R, Okell LC, Dorigatti I, et al. Estimates of the severity of coronavirus disease 2019: a model-based analysis. Lancet Infect Dis. Published online March 30, 2020. DOI:https://doi.org/10.1016/S1473-3099(20)30243-7

9. Paradisi M, Rinaldi G. An empirical estimate of the infection fatality rate of COVID-19 from the first Italian outbreak. Preprint published online May 18, 2020. Available at http://dx.doi.org/10.2139/ssrn.3582811

10. Centers for Disease Control and Prevention. COVID-19 Pandemic Planning Scenarios. Published September 10, 2020. https://www.cdc.gov/coronavirus/2019-ncov/hcp/planning-scenarios.html

11. Ferguson NM, Laydon D, Nedjati-Gilani G, Imai N, Ainslie K, Baguelin M, et al. Impact of non-pharmaceutical interventions (NPIs) to reduce COVID-19 mortality and healthcare demand. Imperial College London. Published online 16 March, 2020. doi: https://doi.org/10.25561/77482

12. Scrucca L, Fop M, Murphy TB and Raftery AE. mclust 5: clustering, classification and density estimation using Gaussian finite mixture models. The R Journal 2016; 8(1):289–317 https://doi.org/10.32614/RJ-2016-021

13. Sylvestre MP, McCusker J, Cole M, Regeasse. Classification of patterns of delirium severity scores over time in an elderly population. International Psychogeriatrics, 2006; 18(4):667–680. doi:10.1017/S1041610206003334.

14. South, A. rworldmap: A New R package for Mapping Global Data. The R Journal 2011; 3(1):35–43.

15. WHO World Population Prospects 2020 estimates. Accessed April 6, 2020. https://population.un.org/wpp/Download/Standard/Population/

16. Our World in Data, https://covid.ourworldindata.org/data/owid-covid-data.csv, extracted on October 13, 2020.

17. Clark A, Jit M, Warren-Gash C, Guthrie B, Wang HHX, Mercer SW, et al. Global, regional, and national estimates of the population at increased risk of severe COVID-19 due to underlying health conditions in 2020: a modelling study. Lancet Glob Health 2020; published online June 12. http://dx.doi.org/10.1016/S2214-109X(20)30264-3.

18. Ghisolfi S, Almås I, Sandefur JC, et al. Predicted COVID-19 fatality rates based on age, sex, comorbidities and health system capacity. BMJ Global Health 2020;5:e003094. doi:10.1136/bmjgh-2020-003094

19. Walker PGT, Whittaker C, Watson OJ, et al. The impact of COVID-19 and strategies for mitigation and suppression in low- and middle-income countries. Science 2020; 369 (6502); 413–422 DOI: 10.1126/science.abc0035

20. Yang J, Zheng Y, Gou X, Pu K, Chen Z, Guo Q, et al. Prevalence of comorbidities and its effects in coronavirus disease 2019 patients: A systematic review and meta-analysis. Int J Infect Dis. 2020;94:91□95. doi:10.1016/j.ijid.2020.03.017

21. Richardson S, Hirsch JS, Narasimhan M, Crawford JM, McGinn T, Davidson KW, et al. Presenting Characteristics, Comorbidities, and Outcomes Among 5700 Patients Hospitalized With COVID-19 in the New York City Area. JAMA. Published online April 22, 2020. doi:10.1001/jama.2020.6775

22. Dowd J, Andriano L, Brazel DM, Rotondi V, Block P, Ding X, et al. Demographic science aids in understanding the spread and fatality rates of COVID-19. Proceedings of the National Academy of Sciences. 2020; 117 (18): 9696–9698; DOI: 10.1073/pnas.2004911117

23. Esteve A, Permanyer I, Boertien Vaupel JW. National age and coresidence patterns shape COVID-19 vulnerability. Proceedings of the National Academy of Sciences. 2020; 117 (28) 16118–16120; DOI: 10.1073/pnas.2008764117

24. RECOVERY Collaborative Group, Horby P, Lim WS, Emberson JR, Mafham M, Bell JL, et al. Dexamethasone in hospitalized patients with COVID-19 - preliminary report. N Engl J Med. 2020. https://doi.org/10.1056/NEJMoa2021436

25. Kubota Y, Shiono T, Kusumoto B, Fujinuma J. Multiple drivers of the COVID-19 spread: The roles of climate, international mobility, and region-specific conditions. PLOS ONE 2020. Published online September 23. https://doi.org/10.1371/journal.pone.0239385

