## Supporting Material text for "Age related clustering in global COVID-19 infection fatality ratios and death trajectories"

**Age standardisation using weights**

Let $R_{j}\left( t \right)$be an event rate in region $j$ at time $t$. Then

$$R_{j}\left( t \right)=\sum_{a} p_{aj}r_{aj}\left( t \right) (1)$$

where $p_{aj}$is the proportion of population $j$ in age group $a$

$$p_{aj}=\frac{pop_{aj}}{pop_{j}}$$

and $r_{aj}\left( t \right)$ is event rate in age group $a$ in region $j$.

Using direct standardisation for region $j$ compared with a reference population, the age adjusted event rate per population is

$$R_{j_{adj}}\left( t \right)=\frac{\sum_{a} {pop}_{a,ref}r_{aj}\left( t \right)}{\sum_{a} {pop}_{a,ref}}$$

$$=\frac{\sum_{a} {pop}_{a,ref}r_{aj}\left( t \right)}{pop_{ref}}$$

$$=\sum_{a} p_{a,ref}r_{aj}\left( t \right)$$

$$=\sum_{a} p_{aj}r_{aj}\left( t \right)*\frac{\sum_{a} p_{a,ref}r_{aj}\left( t \right)}{\sum_{a} p_{aj}r_{aj}\left( t \right)}$$

$$=\frac{R_{j}\left( t \right)}{w_{j,ref}} (2)$$

where $w_{j,ref}$ is defined as the weight for region $j$ compared with the reference population.

$$w_{j,ref}=\frac{\sum_{a} p_{aj}r_{aj}\left( t \right)}{\sum_{a} p_{a,ref}r_{aj}\left( t \right)} (3)$$

**Infection Fatality Ratio**

Let $R_{j}\left( t \right)=IFR_{j}$ be the overall infection fatality ratio in region $j$ and $IFR_{aj}$be the age stratified infection fatality ratio for age group $a$. Age stratified infection fatality ratios from China and other regions have been reported [7-9], so weights for China or Italy relative to other regions can be calculated using Equation (3) and the population age profiles in each region.

$$w_{j,ref}=\frac{\sum_{a} p_{aj}IFR_{aj}}{\sum_{a} p_{a,ref}IFR_{aj}} (4)$$

If rates from China are standardised to other populations, then from Equation (2) and the overall IFR from China, ${IFR}_{China}=0.66\%$ [8], the point estimate for the IFR for China standardised to a reference region $k$ is

$${IFR}_{{China}_{adj}}=\frac{{IFR}_{China}}{w_{China,k}}=\frac{0.66\%}{w_{China,k}} (5)$$

Similarly, lower and upper confidence limits for ${IFR}_{{China}_{adj}}$ can be estimated using the 95% credible interval for ${IFR}_{China}$ and $w_{China,k}$: (lower, upper) =$(\frac{0.39\%}{w_{China,k}},\frac{1.33\%}{w_{China,k}})$

**Death rates per population**

Consider when $R_{j}\left( t \right)$ is a death rate per population in region $j$ at time $t.$

$$R_{j}\left( t \right)=\frac{n_{j}\left( t \right)}{pop_{j}}=\frac{\sum_{a} n_{aj}\left( t \right)}{pop_{j}}=\sum_{a} \frac{{pop_{aj}*n}_{aj}\left( t \right)}{pop_{j}*pop_{aj}}=\sum_{a} p_{aj}r_{aj}\left( t \right)$$

where $r_{aj}\left( t \right)$ is the death rate in age group $a$ in region $j$.

$$r_{aj}\left( t \right)=\frac{n_{aj}\left( t \right)}{pop_{aj}}$$

and $n_{aj}\left( t \right)$ is the number of deaths in age group $a$ in region $j$.

Equation (3) can be re-written as

$$w_{j,ref}=\frac{\sum_{a} p_{aj}\frac{r_{aj}\left( t \right)}{r_{80+,j}}}{\sum_{a} p_{a,ref}\frac{r_{aj}\left( t \right)}{r_{80+,j}\left( t \right)}}$$

$$=\frac{\sum_{a} p_{aj}{r_{rel}}_{a,j}(t)}{\sum_{a} p_{a,ref}{r_{rel}}_{a,j}(t)} (6)$$

where $r_{rel,aj}\left( t \right)=\frac{r_{aj}(t)}{r_{80+,j}(t)}$ are the rates relative to the 80+ age group. If $I_{aj}\left( t \right)=\frac{n_{infection_{a}}\left( t \right)}{pop_{aj}}$ is the infection rate in age group $a$ in region $j$ at time $t$, then

$$r_{aj}\left( t \right)=IFR_{aj}I_{aj}\left( t \right)$$

and

$$r_{rel,aj}\left( t \right)=\frac{IFR_{a}I_{aj}(t)}{IFR_{80+}I_{80+,j}(t)} (7)$$

If the disease spreads evenly through the population, the infection rate is constant across age groups at every time $t$, so that $I_{aj}\left( t \right)=I_{80+,j}(t)$ for all $t$ and the relative event rate is stable over time, ${r_{rel}}_{aj}\left( t \right)={r_{rel}}_{aj}.$ Further, if $\frac{IFR_{aj}\left( t \right)}{IFR_{80+,j}\left( t \right)}$ is intrinsic to the disease (accounting for age but excluding other risk factors such as co-morbidities and obesity) and constant for every region $j$

$$r_{rel,aj}\left( t \right)\approx r_{rel,a}=\frac{IFR_{a}}{IFR_{80+}}$$

Then an approximation to the weight $w_{j,ref}$is

$${w'}_{j,ref}\approx\frac{\sum_{a} p_{aj}{r_{rel}}_{a}}{\sum_{a} p_{a,ref}{r_{rel}}_{a}} (8)$$

To summarise, the weights in Equation (8) are equivalent to Equation (4) if: (1) ${r_{rel}}_{a}$ is the same for every nation and (2) the infection rate is constant across age groups at every time $t$. Estimates of age stratified infection fatality ratios for different regions are presented in Table S1, as well as relative rates ${r_{rel}}_{a}$compared with the 80+ age groups for China and Italy. Although different age groupings are used, the rates are in broad agreement. The exception is the relative rates for 50-60 age group. Since the Chinese study had the highest number of detected cases in the 50–59 age group relative to population size and the Italian study to date is a preprint, data from China were assumed to be more accurate.

In addition,

$${w'}_{j,ref}=\frac{1}{{w'}_{ref,j}}$$

Let region $j$ be the population of interest. The age adjusted death rates per population for the region $j$ standardised to reference region can then be calculated using the crude death rates per population $R_{j}\left( t \right)$ using Equation (2)

$$R_{j_{adj}}\left( t \right)=\frac{R_{j}\left( t \right)}{w_{j,ref}}\approx R_{j}\left( t \right)*{w^{'}}_{ref,j} (9)$$

Using the weights derived from the IFRs in China used in Equation (5),

$$R_{j_{adj}}\left( t \right)\approx R_{j}\left( t \right)*w_{China,j}$$

**S1 Table:** **Comparison of global age specific and relative IFRs.** Confidence limits for IFR/IFR 80+ were estimated using the upper and lower limits for IFR and dividing by the point estimate for IFR 80+ (IFR 81+ for Italy).

| **Age group** | **IFR (%) (95% credible interval)** | **IFR/IFR 80+ (%)* (95% confidence limits)** |
| --- | --- | --- |
| **China (Verity et al) [8]** | | |
| 0-9 | 0.00161 (0.000185, 0.0249) | 0.02 (0.00, 0.32) |
| 10-19 | 0.00695 (0.00149, 0.0502) | 0.09 (0.02, 0.64) |
| 20-29 | 0.0309 (0.0138, 0.0923) | 0.40 (0.18, 1.18) |
| 30-39 | 0.0844 (0.0408, 0.185) | 1.1 (0.52, 2.37) |
| 40-49 | 0.161 (0.0764, 0.323) | 2.1 (0.98, 4.14) |
| 50-59 | 0.595 (0.344, 1.28) | 7.6 (4.41, 16.41) |
| 60-69 | 1.93 (1.11, 3.89) | 24.7 (14.2, 49.9) |
| 70-79 | 4.28 (2.45, 8.44) | 54.9 (31.4, 108.2) |
| 80+ | 7.80 (3.80, 13.3) | 100.0 |
| **Italy (Paradisi) [9]** | | |
| 0-20 | 0.049 (0.0048, 0.178) | 0.54 (0.05, 1.97) |
| 21-40 | 0.0176 (0.0008, 0.0952) | 0.19 (0.01, 1.05) |
| 41-50 | 0.0476 (0.0020, 0.200) | 0.53 (0.02, 2.21) |
| 51-60 | 0.1076 (0.0096, 0.314) | 1.19 (0.11, 3.47) |
| 61-70 | 1.028 (0.588, 1.74) | 11.4 (6.50, 19.3) |
| 71-80 | 4.66 (3.32, 6.99) | 51.6 (36.7, 77.3) |
| 81+ | 9.04 (6.62, 13.3) | 100.0 |
| **Meta-analysis (Levin) [7]** | | |
| 0-34 | 0.004 (0.003,0.005) | - |
| 35-44 | 0.068 (0.058, 0.078) | - |
| 45-54 | 0.23 (0.20, 0.26) | - |
| 65-74 | 2.5 (2.1, 3.0) | - |
| 75-84 | 8.5 (6.9, 10.4) | - |
| 85+ | 28.3 (21.8, 36.6) | - |

*IFR/IFR 81+ (%) for Italy; not calculated for meta-analysis since the highest age bracket was 85+

**Model diagnostics for clustering of Infection Fatality Ratios**

S1 Fig shows the probability density function from the sum of the three Gaussian normal distribution and the observed histogram. Tests for normality (Quantile-Quantile plot) and model fit are found in S2 Fig and provide evidence of good model fit.

**S1 Figure**: **Probability density function and histogram.** Probability density function of fitted distributions from three clusters (solid line) and histogram of observed IFRs.

**S2 Figure:** **Model diagnostics.** Quantile-Quantile plots to test normality (left) and estimated and empirical cumulative density functions (CDF) (right).

**Trajectory analysis**

The ‘traj’ package used three steps to classify trajectories into clusters: (1) calculating 24 measures describing the change in death rates over time (eg direction of change, nonlinearity, fluctuations) [https://cran.r-project.org/web/packages/traj/vignettes/trajVignette.pdf]; (2) A subset of the measures which describe the main features of the trajectories was selected using factor analysis and (3) the selected measures were clustered with the R package ‘Nbclust’ to determine the optimal number of clusters, using k-means clustering with 2 to 15 clusters and selecting the number of clusters based on the maximum cubic clustering criterion (ccc).

Statistical measures selected using principal components analysis and associated eigenvalues can be found in S2 Table. Two clusters were found for 50 day (S3 Fig) and three clusters for 150 day (S4 Fig) trajectories. There is evidence that the trajectories are clustered into more than one group, because the within groups sum of squares in the scree plots for two/three clusters is lower than for one cluster (S3 and S4 Figs).

**S2 Table: Statistical measures of trajectory clusters.** Statistical measures selected by factor analysis to describe the 50 and 150 day trajectories by cluster.

| **Statistical Measure** | **Description** | **Eigenvalue** | **Cluster 1 (‘Steady’) median (IQR)** | **Cluster 2 (‘Moderate’) median (IQR)** | **Cluster 3 (‘Fast’) median (IQR)** |
| --- | --- | --- | --- | --- | --- |
| **50 day trajectory** | | | | |  |
| Measure 1 | Change relative to the mean over time | 8.25 | 2.49 (2.09-2.68) | 3.90 (3.51-4.36) | - |
| Measure 2 | Mean of the absolute second differences | 5.37 | 0.01 (0.00-0.02) | 0.02 (0.00-0.07) | - |
| Measure 3 | Ratio of the maximum absolute second difference to mean absolute first difference | 1.64 | 0.18 (0.12-0.25) | 0.17 (0.13-0.25) | - |
| **150 day trajectory** | | | | |  |
| Measure 1 | Standard Deviation (SD) | 7.71 | 118.9 (110.7-173.5) | 24.4 (13.0-64.2) | 9.44 (3.73-17.1) |
| Measure 2 | Coefficient of variation | 4.79 | 80.0 (64.0-107.9) | 105.8 (95.3-128.3) | 57.0 (49.3-69.6) |
| Measure 3 | Ratio of the SD of the  first differences to the slope | 2.88 | 0.93 (0.67-1.03) | 0.87 (0.69-1.27) | 0.87 (0.63-1.15) |
| Measure 4 | Ratio of the maximum absolute second difference to mean absolute first difference | 1.30 | 0.22 (0.13-0.29) | 0.17 (0.11-0.26) | 0.24 (0.18-0.35) |

**S3 Figure: Criteria used to determine optimal number of clusters for 50 day trajectories.** Cubic clustering criterion (ccc) for 2-15 clusters (left) and within groups sum of squares for 1-15 clusters (right).

**S4 Figure: Criteria used to determine optimal number of clusters for 150 day trajectories.** Cubic clustering criterion (ccc) criteria for 2-15 clusters (left) and within groups sum of squares for 1-15 clusters (right).
