## Supplementary figures and images for "Age related clustering in global COVID-19 infection fatality ratios and death trajectories"

### S1 Fig

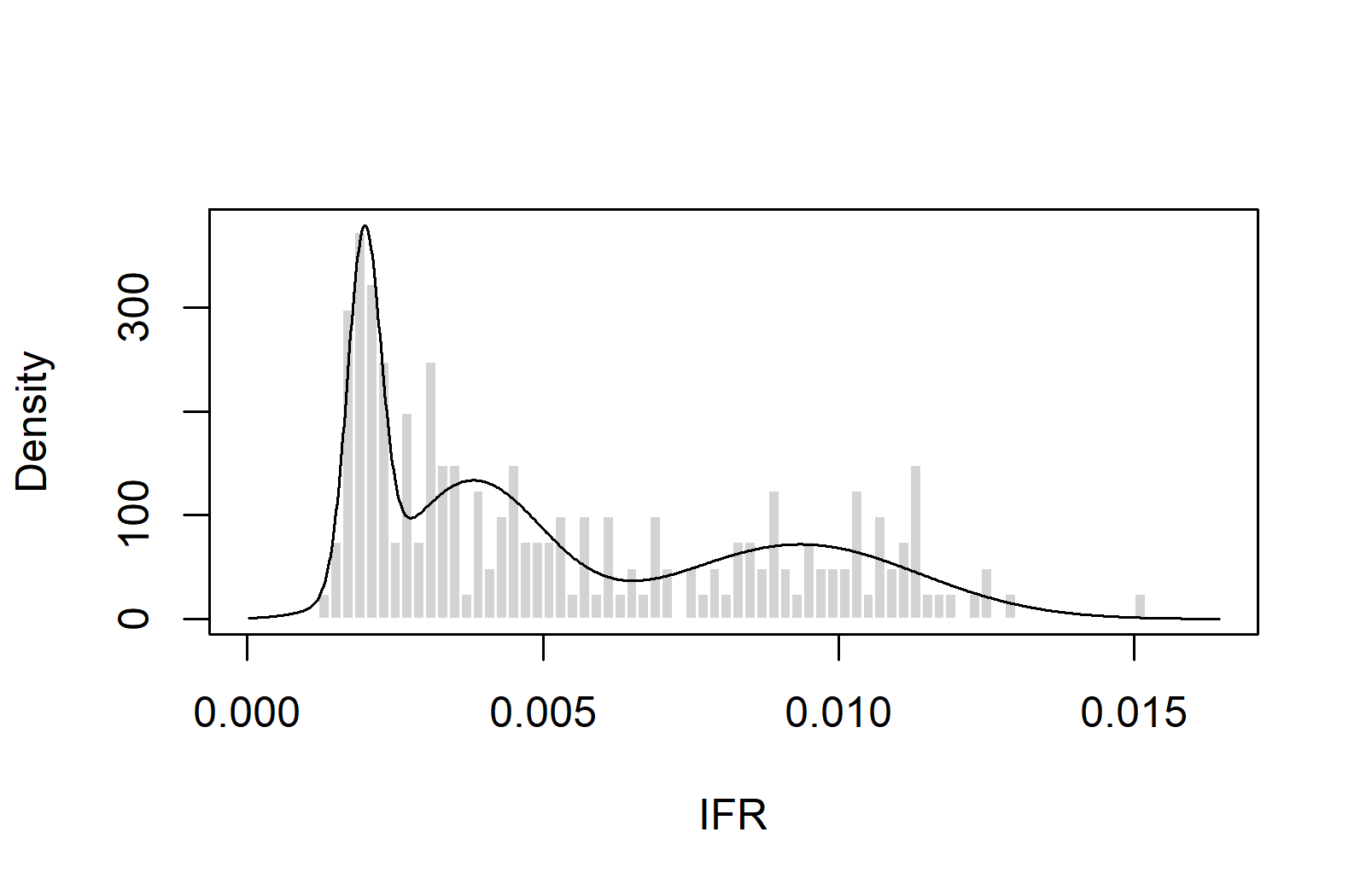

### S2 Fig

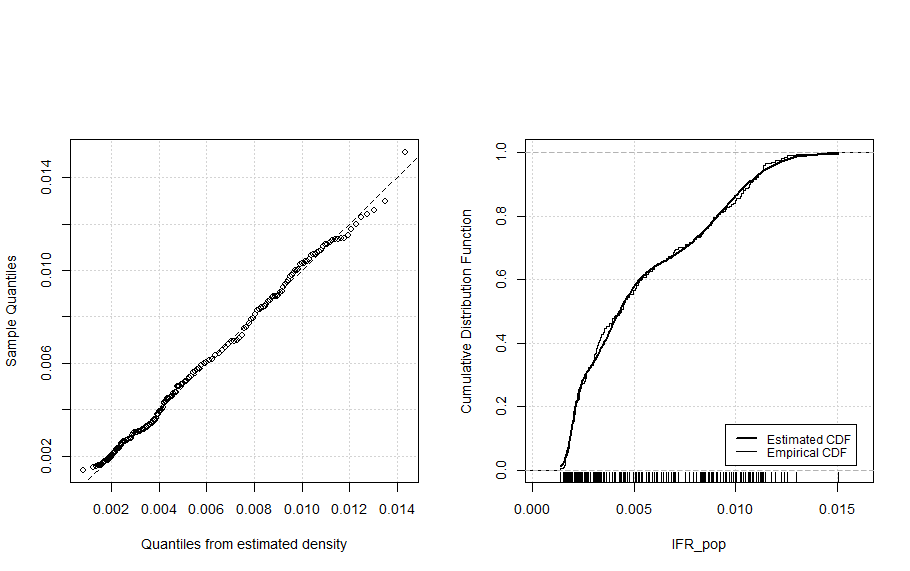

### S3 Fig

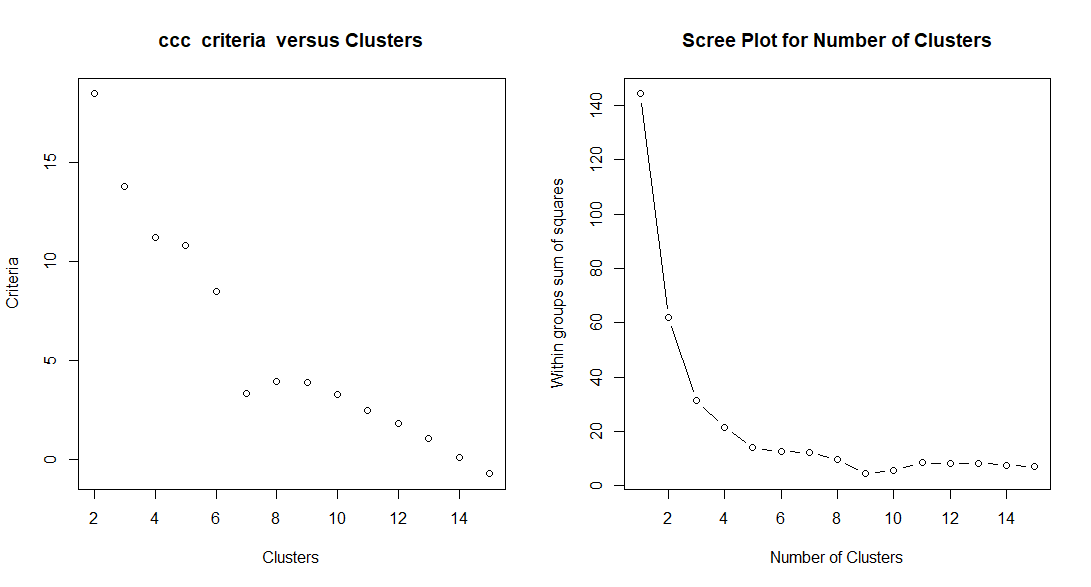

### S4 Fig

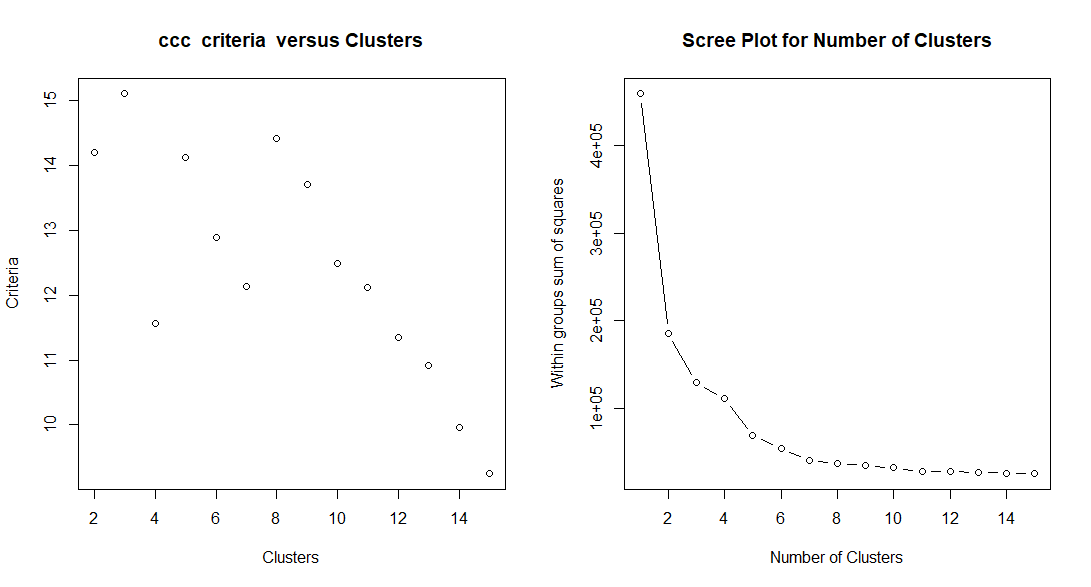
